# Multicenter Development and Prospective Validation of eCARTv5: A Gradient Boosted Machine Learning Early Warning Score

**DOI:** 10.1101/2024.03.18.24304462

**Authors:** Matthew M. Churpek, Kyle A. Carey, Ashley Snyder, Christopher J Winslow, Emily Gilbert, Nirav S Shah, Brian W. Patterson, Majid Afshar, Alan Weiss, Devendra N. Amin, Deborah J. Rhodes, Dana P. Edelson

**Author notes:** Corresponding author: Matthew M Churpek, MD, MPH, PhD.

## Abstract

**OBJECTIVE:** Early detection of clinical deterioration using machine learning early warning scores may improve outcomes. However, most implemented scores were developed using logistic regression, only underwent retrospective validation, and were not tested in important subgroups. Our objective was to develop and prospectively validate a gradient boosted machine model (eCARTv5) for identifying clinical deterioration on the wards.

**DESIGN:** Multicenter retrospective and prospective observational study.

**SETTING:** Inpatient admissions to the medical-surgical wards at seven hospitals in three health systems for model development (2006-2022) and at 21 hospitals from three health systems for retrospective (2009-2023) and prospective (2023-2024) external validation.

**PATIENTS:** All adult patients hospitalized at each participating health system during the study years.

**INTERVENTIONS:** None

**MEASUREMENTS AND MAIN RESULTS:** Predictor variables (demographics, vital signs, documentation, and laboratory values) were used in a gradient boosted trees algorithm to predict intensive care unit transfer or death in the next 24 hours. The developed model (eCART) was compared to the Modified Early Warning Score (MEWS) and the National Early Warning Score (NEWS) using the area under the receiver operating characteristic curve (AUROC). The development cohort included 901,491 admissions, the retrospective validation cohort included 1,769,461 admissions, and the prospective validation cohort included 205,946 admissions. In retrospective validation, eCART had the highest AUROC (0.835; 95%CI 0.834, 0.835), followed by NEWS (0.766 (95%CI 0.766, 0.767)), and MEWS (0.704 (95%CI 0.703, 0.704)). eCART’s performance remained high (AUROC ≥0.80) across a range of patient demographics, clinical conditions, and during prospective validation.

**CONCLUSIONS:** We developed eCART, which performed better than the NEWS and MEWS retrospectively, prospectively, and across a range of subgroups. These results served as the foundation for Food and Drug Administration clearance for its use in identifying deterioration in hospitalized ward patients.

**KEY POINTS:** *Question:* How can we best identify deterioration in hospitalized ward patients?

*Findings:* In retrospective validation, a gradient boosted machine model (eCARTv5) developed for identifying clinical deterioration on the wards had the highest area under the receiver operating characteristic curve when compared to the Modified Early Warning Score and the National Early Warning Score. eCART’s performance remained high across a range of patient demographics, clinical conditions, and during prospective validation.

*Meaning:* This paper evaluating eCART’s performance served as the foundation for Food and Drug Administration clearance for its use in identifying deterioration in hospitalized ward patients.

## INTRODUCTION

Clinical deterioration occurs in up to 5% of hospitalized patients, and early identification is associated with improved outcomes (1–9). These events are often heralded by deranged physiology, leading to the development of early warning scores aimed at identifying high-risk patients (10, 11). Early warning scores have evolved from aggregated weighted scores, such as the Modified Early Warning Score (MEWS) (12), which can be calculated by hand, to those based on logistic regression (1, 2), which can be summed with a calculator or spreadsheet, and more recently, to advanced machine learning models, such as gradient boosted machines (GBM), which can be more accurate in large datasets (13–16).

While machine learning models may decrease false alarms and increase detection rates, they can suffer from poor performance when trained on insufficiently sized or representative datasets (17, 18). Furthermore, their use in clinical practice raises concerns about model fairness and bias, and little is known regarding how they perform across a range of important patient subgroups. Evaluating models across subgroups, such as age, sex, and race, is critical to ensuring the fairness of these tools in practice so that they have the potential to benefit all patients. Finally, most prior work has been done retrospectively (11), and it is not known whether these complex models will perform similarly in prospective production environments.

Therefore, we aimed to develop and externally validate a GBM model for identifying clinical deterioration in a large, geographically diverse set of hospitals. After retrospective validation, which included extensive subgroup analyses, we tested performance prospectively. These results served as the foundation for 510(k) clearance by the U.S. Food and Drug Administration (FDA) for inpatient clinical deterioration detection outside the intensive care unit (ICU) (19).

## METHODS

### Study Setting and Population

Hospitalized adults (age ≥18 years) admitted to the medical-surgical wards were eligible for inclusion in this observational cohort study. Patients who were only admitted to the ICU, labor and delivery, or emergency department and were never transferred to a medical-surgical (non-ICU) unit during their hospital encounter were excluded. The model, eCARTv5 (eCART), was developed in a dataset of seven hospitals from three health systems in Illinois (D1-D3), spanning 2006-2022. The model was then externally validated in two phases: (1) a retrospective cohort of admissions to 21 hospitals from three health systems (R1-R3) in Florida, Wisconsin, Connecticut, and Rhode Island (2009-2023); and (2) a prospective cohort of consecutive admissions to the same hospitals (P1-P3). eCART was compared to the MEWS (12) and the National Early Warning Score (NEWS) (20), commonly used tools for clinical deterioration. The original NEWS was chosen as a comparator over NEWS2 because choosing the SpO2 scale in NEWS2 requires real-time clinician determination (21). The study was approved by the following Institutional Review Boards (IRB): University of Chicago Biological Sciences Division IRB (#18-0447), Loyola University Chicago Health Sciences Division IRB (#215437), NorthShore University HealthSystem Research Institute IRB (#EH16-210T), University of Wisconsin-Madison Minimal Risk Research IRB (#2019-1258), BayCare Health System IRB (#2022.014-B.MPH; 2022.015-B.MPH), and Yale Human Research Protection Program IRBs (#2000035317). Each IRB waived study-specific informed consent. Procedures followed the ethical standards of the responsible institutional committee on human experimentation and the Helsinki Declaration of 1975.

### Main Outcome Measure

The study outcome was clinical deterioration, defined as a ward death or ICU transfer from the wards within 24 hours of a score (1, 13, 20). Death was determined using the discharge disposition in the EHR, with the time of death being the last recorded vital sign. ICU transfer was defined as a direct ward-to-ICU transfer and was determined using the transfer disposition in the EHR.

### Predictor Variables

Ninety-seven features were included as predictor variables in the eCART model. These variables included patient characteristics (e.g., age, body mass index (BMI)), vital signs, laboratory values, time of day, and nursing/respiratory therapist documentation (e.g., delivered oxygen percentage, Braden scale), as well as trends (22). A full list of model predictor variables is found in Appendix Table E1 in the Online Supplement.

Non-physiologic flowsheet data were considered input errors and treated as missing, as per prior publications (see Appendix Table E2 in the Online Supplement). If no prior values were available at a specific time, then the most recent prior value, if available, was pulled forward. If no prior values were available, the variable was left as missing. Given the dynamic nature of blood gas and lactate values and the tendency of providers to only order them on actively deteriorating patients, those values were only pulled forward for 24 hours, after which they were treated as missing.

### Model Development

A GBM model was developed to predict clinical deterioration in the training data using discrete-time survival analysis (1, 13, 23). Time was discretized into 8-hour blocks, and the data at the beginning of each segment were used to predict whether an outcome occurred within the following eight hours. This approach allowed the inclusion of time-varying predictors, removed the bias of sicker patients receiving more frequent measurements, and provided results analogous to a Cox survival model (23). Because tree-based models can perform poorly in imbalanced data (i.e., when the outcome of interest is uncommon), down-sampling of the training dataset to obtain a 50% outcome prevalence was performed before model fitting (13). Model hyperparameters were tuned in the training cohort using five-fold cross-validation to maximize the area under the receiver operating characteristic curve (AUROC). No variable selection was performed, as research has shown that this may degrade the accuracy of tree-based algorithms (24). Variable importance was calculated based on the improvement of each split averaged across all trees (17). Additional details can be found in the Online Supplement.

### Retrospective and Prospective Score Calculation

For model validation, data pre-processing was performed in the same manner as during model development, except that in the validation cohorts, data were not blocked, and no down-sampling was performed. Specifically, each time a new observation was recorded in the EHR, predicted probabilities from the eCART model, MEWS, and NEWS values were calculated. The transformed eCART model output probabilities were then scaled to eCART scores ranging from 0-100 for ease of interpretation. For the retrospective validation, these scores were calculated on a static multicenter dataset stored on secured laboratory servers. In the prospective validation, the model features, eCART scores, and outcomes were collected in real-time utilizing Health Level-Seven Version 2 (HL-7 V2) messaging standard interfaces, a clinical data standard to protocolize how data are shared and exchanged in EHR operations, and stored on cloud-based computing and storage resources hosted at Amazon Web Services. The engine calculated scores as data became available, with the first score filed back to the EHR for any given time. For the prospective cohort, MEWS and NEWS scores were not calculated in real-time. In a subset of encounters from the prospective cohort, retrospective eCART scores were also calculated to directly compare the impact of the different methodologies on scoring.

### Statistical Analysis

Descriptive statistics were used to characterize patient demographics across the separate development, retrospective validation, and prospective validation cohorts. Model performance was calculated by assessing the ability of the scores at each observation time to predict clinical deterioration in the following 24 hours. Discrimination was measured using the AUROC and then compared using DeLong’s method (25). To demonstrate that the inclusion of multiple observations per encounter did not bias the results, a 100-iteration cluster bootstrapped analysis with replacement was run in the retrospective and prospective cohorts (26, 27). Subgroup analyses were performed in the retrospective validation cohort across patient demographics and clinical conditions (see Supplementary Methods in the Online Supplement for definitions). Sensitivity, specificity, and positive and negative predictive values were calculated for each threshold, with confidence intervals calculated using the Clopper-Pearson method. Performance at a moderate-risk and high-risk threshold for each score (eCART of ≥93 and ≥97; NEWS ≥5 and ≥7; MEWS ≥3 and ≥4) was also compared. Model calibration was assessed in the prospective validation by comparing observed to expected deterioration rates across eCART score values. Analyses were performed using Stata version 16.1 (StataCorps; College Station, Texas) and R version 4.2.1 (The R Foundation for Statistical Computing, Vienna, Austria).

## RESULTS

The training dataset included 901,491 adult inpatient admissions with ward stays at seven hospitals from three health systems. The validation cohorts included 1,769,461 retrospective admissions and 205,946 non-overlapping prospective admissions to 21 hospitals from three health systems. There was variation in demographics across the three cohorts (Table 1). When compared to the two validation cohorts, the training cohort had a higher proportion of black patients (32% vs. 14%) and a lower percentage of sepsis, COVID-19, heart failure, and COPD. Meanwhile, the prospective validation cohort differed from the retrospective validation cohort in having a higher median age (66 vs. 62 years), a shorter length of stay (60 vs. 72 hours), and fewer encounters with surgical procedures. There was also variation in missing variables across the health systems (Table E3 and E4 in the Online Supplement). Most notably, R3 had higher rates of missing Braden scores and respiratory rate trends, suggesting a lower frequency of respiratory rate documentation, which was more pronounced in the prospective cohort (P3). R1 had higher rates of missing hematology labs, particularly white blood cell differential distributions. R2 had higher missing rates for mental status.

**Table 1.** Patient characteristics by cohort. Abbreviations: ICU = Intensive Care Unit; COVID-19 = Coronavirus Disease 2019; AIDS/HIV = Human Immunodeficiency Virus/Acquired Immunodeficiency Syndrome

| Patient Characteristics | Derivation<br>Cohort | Retrospective<br>Validation Cohort | Prospective<br>Validation Cohort |
| --- | --- | --- | --- |
| Hospitals, N | 7 | 21 | 21 |
| Encounters, N | 901,491 | 1,769,461 | 205,946 |
| Admission age, years, median (IQR) | 61 (44, 74) | 62 (45, 75) | 66 (52, 78) |
| Female sex | 512,198 (56.8%) | 993,297 (56.1%) | 113,422 (55.1%) |
| Race: American Indian or Alaska Native | 1,445 (0.2%) | 6,468 (0.4%) | 538 (0.3%) |
| Race: Asian/Mideast Indian | 23,334 (2.6%) | 26,681 (1.5%) | 2,928 (1.4%) |
| Race: Black/African American | 286,901 (31.8%) | 252,982 (14.3%) | 28,659 (13.9%) |
| Race: Pacific Islander/ Hawaiian Native | 666 (0.1%) | 2,496 (0.1%) | 224 (0.1%) |
| Race: White/Caucasian | 496,154 (55.0%) | 1,384,075 (78.2%) | 158,435 (76.9%) |
| Race: Other | 92,991 (10.3%) | 96,759 (5.5%) | 15,162 (7.4%) |
| Surgical | 312,153 (34.6%) | 539,875 (30.5%) | 49,265 (23.9%) |
| Obstetric | 65,594 (7.3%) | 154,759 (8.7%) | 12,653 (6.1%) |
| Sepsis | 240,651 (26.7%) | 639,802 (36.2%) | 77,296 (37.5%) |
| COVID-19 | 4,365 (0.5%) | 49,834 (2.8%) | 7,920 (3.8%) |
| Congestive heart failure | 131,644 (14.6%) | 306,140 (17.3%) | 44,223 (21.5%) |
| Chronic pulmonary disease | 150,043 (16.6%) | 443,263 (25.1%) | 53,820 (26.1%) |
| Length of stay, hours, median (IQR) | 70 (37, 124) | 72 (43, 130) | 65 (35, 116) |
| Ward to ICU transfer | 32,320 (3.6%) | 57,789 (3.3%) | 6,143 (3.0%) |
| Mortality | 10,568 (1.2%) | 26,319 (1.5%) | 2,373 (1.2%) |

The most important variables in the final eCART model were maximum respiratory rate in the prior 24 hours, delivered FiO2, minimum systolic blood pressure in the prior 24 hours, and heart rate (Figure 1). Partial plots illustrating the relationship between the values of these variables and the risk of deterioration are shown in Figure E1.

**Figure 1.**
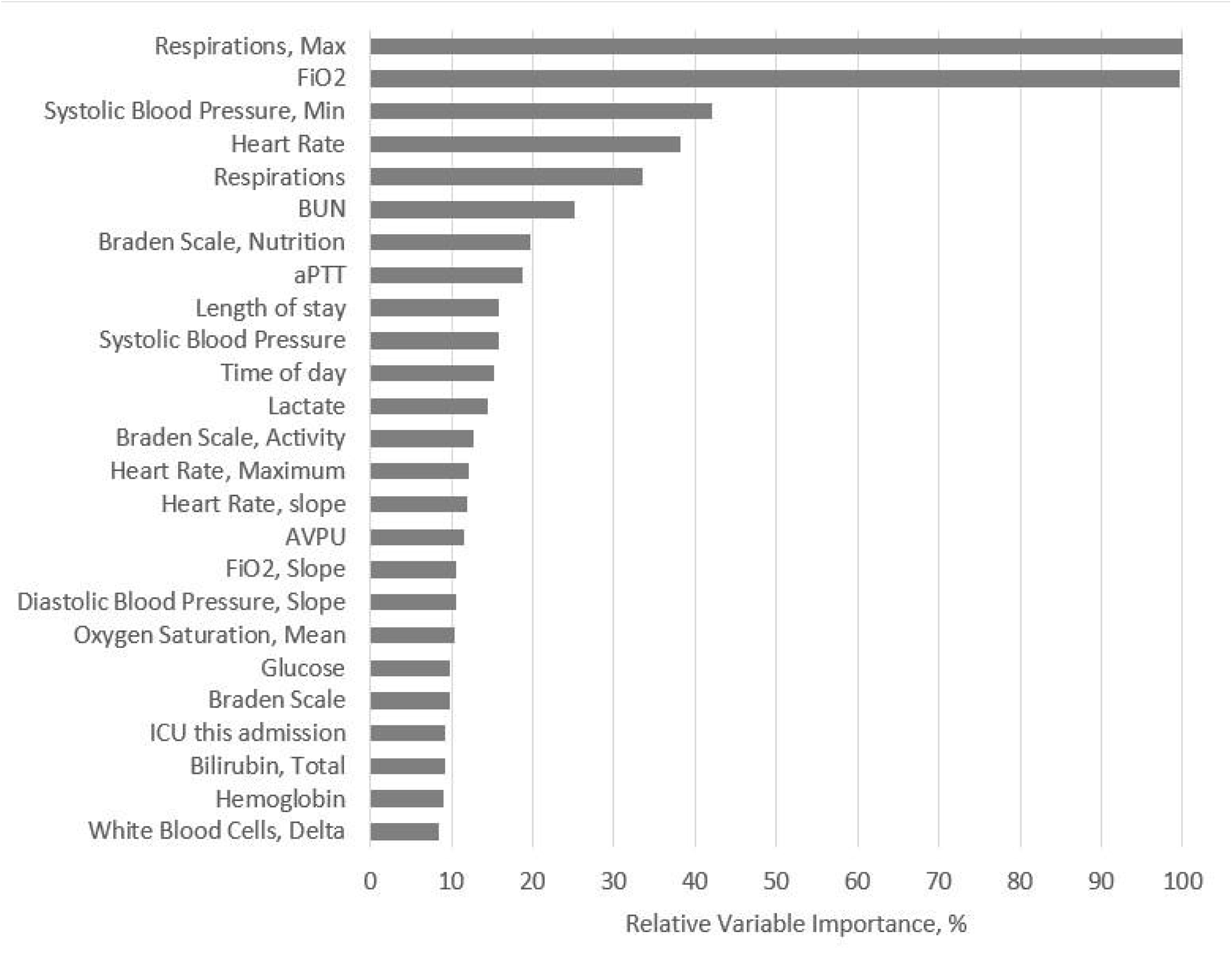
Variable importance plot illustrating the top 25 most important variables in the eCART model. Abbreviations: eCART = electronic Cardiac Arrest Risk Triage score; FiO2 = Fraction of Inspired Oxygen; BUN = Blood Urea Nitrogen; aPTT = Activated Partial Thromboplastin Clotting Time; AVPU = Alert, responds to Voice, responds to Pain, Unresponsive

In the retrospective validation dataset, 132,873,833 eCART, MEWS, and NEWS scores were calculated. The AUROC for eCART was 0.835 (0.834, 0.835) for the full retrospective cohort (Table 2). eCART consistently outperformed NEWS (AUROC 0.766 (0.766, 0.767)), which consistently outperformed MEWS (AUROC 0.704 (0.703, 0.704)). eCART’s sensitivity in the retrospective cohort at the moderate-risk threshold (≥93) was 51.8%, with a positive predictive value (PPV) of 9.0%. At the high-risk threshold (≥97), PPV increased to 14.2% with a decrease in sensitivity to 38.6% (Appendix Table E5 in the Online Supplement). In contrast, MEWS had sensitivities of 38.9% and 22.6% at the moderate (≥3) and high-risk (≥4) thresholds, with corresponding PPVs of 5.7% and 10.4%, respectively, while NEWS had sensitivities of 49.7% and 28.0% at the moderate (≥5) and high-risk (≥7) thresholds, with corresponding PPVs of 5.4% and 9.9%, respectively (Appendix Tables E6 and E7 in the Online Supplement). The precision-recall curve, Figure 2, plots PPV as a function of sensitivity and demonstrates a consistently more favorable tradeoff between sensitivity and PPV for eCART compared to both NEWS and MEWS. At the moderate-risk threshold, NEWS provided the longest lead time before the deterioration event (17 hours (IQR 1, 73)), followed by eCART (16 hours (IQR 1, 68)), and then MEWS (13 hours (IQR 1, 66)). At the high-risk threshold, eCART alerted a median of 5 (IQR 0, 43) hours in advance of clinical deterioration, significantly earlier than NEWS (3 hours (IQR 0, 40) and MEWS (2 hours (IQR 0, 30)) (p<0.01 for all comparisons).

**Figure 2.**
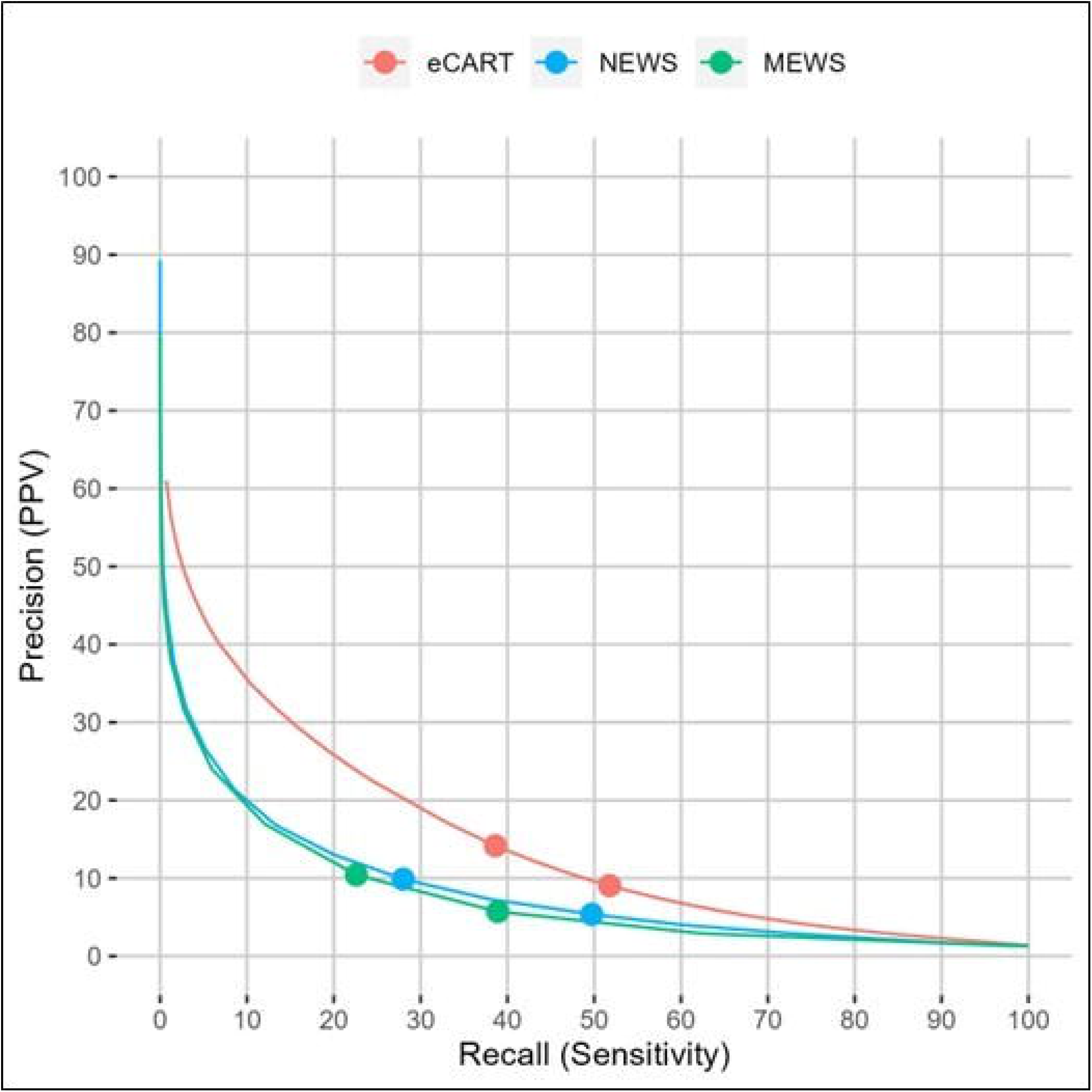
Precision-recall curves of the risk scores in the full retrospective dataset (n=132,873,833 observations). Sensitivity is plotted along the X-axis and positive predictive value is plotted along the Y-axis for eCART, NEWS and MEWS. The markers on the lines correspond to a MEWS of 3 and 4, NEWS of 5 and 7 and eCART of 93 and 97, representing commonly used moderate (higher sensitivity) and high-risk (higher PPV) thresholds for each score. Abbreviations: eCART = electronic Cardiac Arrest Risk Triage score; NEWS = National Early Warning Score; MEWS = Modified Early Warning Score; PPV = Positive Predictive Value

**Table 2.** Area under the receiver operating characteristic curve (AUROC) of the risk scores for predicting the outcome of clinical deterioration within 24 hours in the external retrospective validation cohort. Abbreviations: eCART = electronic Cardiac Arrest Risk Triage score; NEWS = National Early Warning Score; MEWS = Modified Early Warning Score

| Cohort | Encounters,<br>n | Observations,<br>n | eCART<br>AUROC (95% CI) | NEWS<br>AUROC (95% CI) | MEWS<br>AUROC (95% CI) |
| --- | --- | --- | --- | --- | --- |
| **Retrospective (All)** | 1,769,461 | 132,873,833 | 0.835 (0.834,<br>0.835) | 0.766 (0.766,<br>0.767) | 0.704 (0.703,<br>0.704) |
| •**Retrospective (R1)** | 246,949 | 19,262,093 | 0.862 (0.861,<br>0.862) | 0.775 (0.774,<br>0.777) | 0.730 (0.729,<br>0.732) |
| •**Retrospective (R2)** | 592,504 | 37,930,348 | 0.872 (0.872,<br>0.873) | 0.808 (0.807,<br>0.809) | 0.749 (0.748,<br>0.749) |
| •**Retrospective (R3)** | 930,008 | 75,681,392 | 0.807 (0.807,<br>0.808) | 0.744 (0.743,<br>0.744) | 0.674 (0.674,<br>0.675) |
| **Prospective (All)** | 205,946 | 21,516,964 | 0.828 (0.827,<br>0.829) | 0.767 (0.766,<br>0.768) | 0.709 (0.708,<br>0.710) |
| •**Prospective (P1)** | 8,036 | 1,270,931 | 0.855 (0.851,<br>0.858) | 0.768 (0.764,<br>0.773) | 0.735 (0.731,<br>0.739) |
| •**Prospective (P2)** | 33,092 | 2,929,075 | 0.874 (0.872,<br>0.875) | 0.809 (0.807,<br>0.811) | 0.757 (0.755,<br>0.760) |
| •**Prospective (P3)** | 164,818 | 17,316,958 | 0.817 (0.816,<br>0.818) | 0.758 (0.757,<br>0.759) | 0.697 (0.696,<br>0.698) |

Performance across subgroups in the retrospective validation (Table 3) demonstrated that eCART (AUROCs 0.810-0.909) consistently outperformed NEWS (AUROCs 0.745-0.793), which outperformed MEWS (AUROCs 0.672-0.726). Among the different age groups, the AUROC for eCART was highest in 18-33 year-old patients (0.861) and lowest in 65-78 year-old patients (0.822). In subgroup analysis by race, the eCART AUROC was highest in the Native Hawaiian/Other Pacific Islander cohort (0.862) and lowest in the American Indian or Alaska Native cohort (0.814). eCART performance was slightly higher for female compared to male patients (0.844 vs 0.824) and was exceptionally high in obstetric encounters (0.909). Among the clinical conditions, performance was highest in patients with COVID-19 across all scores and lowest in heart failure. Across all subgroups studied, eCART retained high discrimination (AUROC ≥0.81).

**Table 3.** Subgroup analysis results showing the area under the receiver operating characteristic curve (AUROC) values for predicting clinical deterioration in the full retrospective cohort by risk score and subgroup. Abbreviations: eCART = electronic Cardiac Arrest Risk Triage score; NEWS = National Early Warning Score; MEWS = Modified Early Warning Score; COVID-19 = Coronavirus Disease 2019; CHF = Congestive Heart Failure; COPD = Chronic Obstructive Pulmonary Disease

| Category | Subgroup | Encounters, n | eCART<br>AUROC | NEWS<br>AUROC | MEWS<br>AUROC |
| --- | --- | --- | --- | --- | --- |
| All | — | 1,769,461 | 0.835 | 0.766 | 0.704 |
| Age | 18-33 | 232,353 | 0.861 | 0.770 | 0.726 |
|  | 34-48 | 271,904 | 0.845 | 0.758 | 0.709 |
|  | 49-64 | 475,033 | 0.827 | 0.755 | 0.702 |
|  | 65-78 | 458,470 | 0.822 | 0.758 | 0.700 |
|  | ≥79 | 331,701 | 0.832 | 0.776 | 0.716 |
| Sex | Male | 776,164 | 0.824 | 0.761 | 0.699 |
|  | Female | 993,297 | 0.844 | 0.775 | 0.710 |
| Race | American Indian or Alaska Native | 6,468 | 0.814 | 0.746 | 0.672 |
|  | Asian/Mideast Indian | 26,681 | 0.847 | 0.779 | 0.722 |
|  | Black/African-American | 252,982 | 0.831 | 0.769 | 0.707 |
|  | Native Hawaiian/Other Pacific Islander | 2,496 | 0.862 | 0.772 | 0.709 |
|  | White | 1,384,075 | 0.834 | 0.764 | 0.702 |
|  | Other/Unknown | 96,759 | 0.859 | 0.785 | 0.724 |
| Procedure | Surgical | 539,875 | 0.817 | 0.745 | 0.690 |
|  | Obstetric | 154,759 | 0.909 | 0.758 | 0.691 |
| Clinical condition | Sepsis | 639,802 | 0.836 | 0.774 | 0.714 |
|  | COVID-19 | 49,834 | 0.858 | 0.793 | 0.710 |
|  | CHF | 306,140 | 0.810 | 0.750 | 0.694 |
|  | COPD | 443,263 | 0.824 | 0.757 | 0.698 |

In the prospective analysis, there were 21,516,964 scores calculated, and 7,303 encounters had clinical deterioration within 24 hours following an observation. Performance in the full prospective cohort was similar to the full retrospective cohort, with eCART (AUROC 0.828) outperforming both NEWS (AUROC 0.767) and MEWS (AUROC 0.709). Model calibration is shown in Figure 3, demonstrating close agreement between the observed and expected deterioration rates. The bootstrapped analysis resulted in near perfect agreement between the unadjusted and adjusted results for eCART in both the retrospective and prospective cohorts (data not shown). Finally, in a head-to-head comparison of the real-time device output scoring and the retrospectively calculated scores in a convenience sample of 162,335 prospective cohort encounters from P3, the device calculated 17,126,259 eCART scores in real-time in those encounters with a mean value of 53 (SD 28), while the retrospective extract generated 15,251,520 eCART scores with a mean of 49 (SD 29). The resulting AUROCs were slightly higher using the retrospective calculation methodology compared to the prospectively collected score [0.825 (0.824, 0.826) vs 0.817 (0.816, 0.818)].

**Figure 3.**
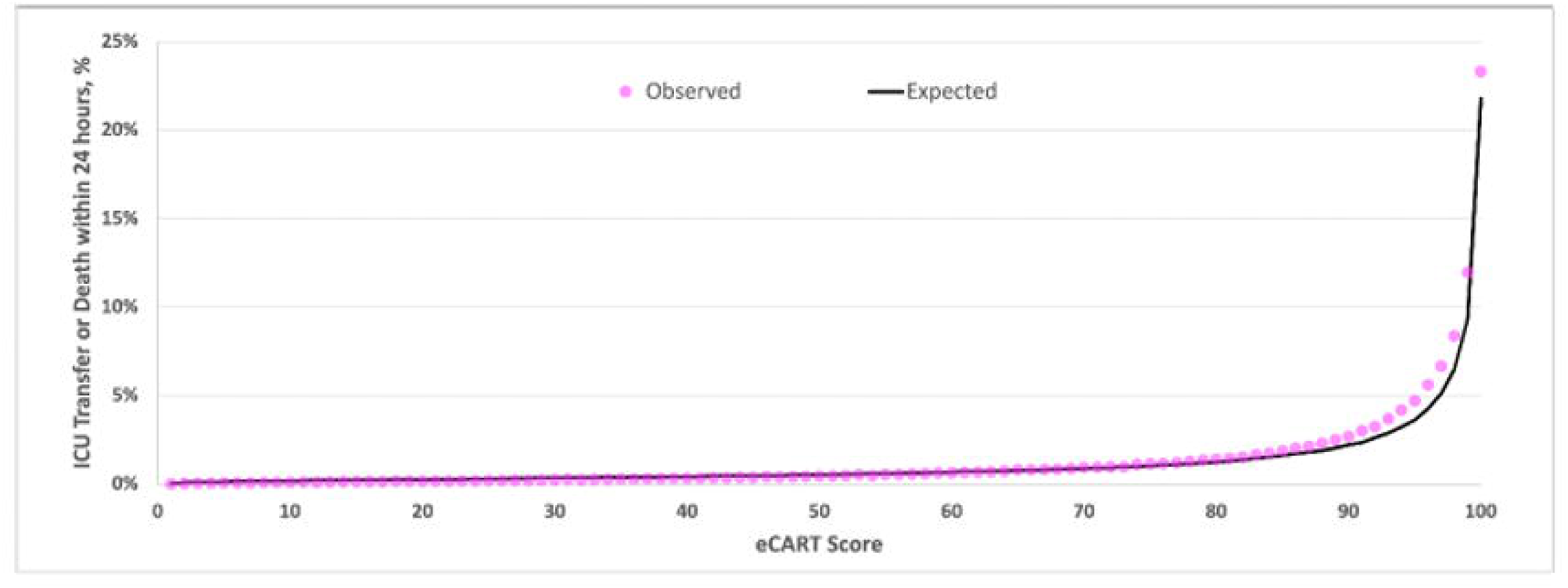
Calibration plot in the prospective validation cohort illustrating the observed and expected outcome rates across values of the eCART score. Abbreviations: eCART = electronic Cardiac Arrest Risk Triage score; ICU = Intensive Care Unit

## DISCUSSION

In a large retrospective validation of nearly two million inpatient encounters with over 130 million calculated scores from three geographically distinct health systems in the United States, eCART outperformed MEWS and NEWS for predicting clinical deterioration. Results were robust across age, sex, and race, with eCART performing better than NEWS and MEWS in all subgroups. Further, eCART had consistently high discrimination in patients with heart failure, COPD, sepsis, and COVID-19, as well as in surgical and obstetric patients. This illustrates an important strength of developing an all-cause deterioration model because it enhances early identification across a wide range of patients instead of more narrowly developed models for specific conditions (e.g., sepsis). Performance was confirmed in a prospective study of over 200,000 admissions. These results, which constitute the largest validation of an early warning score to date, provide confidence that eCART’s performance is strong and generalizable. Prospective implementation of eCART would lead to increased detection and decreased false alarm rates compared to MEWS and NEWS, which could improve patient outcomes and decrease alarm fatigue.

The original version of eCART, which our group developed in 2014, included 269,999 admissions from three health systems in Illinois and used logistic regression (1). The AAM score developed by Kipnis and colleagues used a similar approach to predict unplanned ICU transfer in the Kaiser Permanente Northern California health system (2). Prospective implementation studies of both scores found an association with decreased mortality (8, 9). However, our group and others have found that more advanced machine learning methods, such as GBM, can outperform logistic regression for predicting clinical deterioration (13, 14). Further, including trends has also been shown to improve model discrimination (22). Therefore, in this new version of eCART, we used GBM with trends to optimize performance. The high performance of eCART is consistent with prior early warning scores developed using machine learning, including the Hospital-wide Alerting Via Electronic Noticeboard (HAVEN), which is a GBM model developed and then validated in four hospitals in the United Kingdom that outperformed other scoring systems based on logistic regression (14). The increased discrimination of these more advanced models can increase detection rates while limiting false positive alerts that can cause alarm fatigue.

While previously published studies have demonstrated that more advanced models can outperform standard tools, such as MEWS and NEWS, across the entire medical-surgical cohort (2, 13, 14), little is known regarding comparative performance across important patient subgroups. Therefore, in this study, we performed extensive subgroup analysis that included age, sex, race, and medical conditions. We found that eCART had high discrimination across all subgroups and consistently outperformed both MEWS and NEWS. The highest performance was in the post-partum cohort, followed by age 18-33, Native Hawaiian and Other Pacific Islanders, and patients with COVID-19. Across all 19 tested subgroups, eCART maintained an AUC of ≥0.81. To our knowledge, this is the largest and most comprehensive subgroup analysis performed on early warning scores to date and demonstrates that eCART can be used across a wide range of hospitalized patients.

Although numerous predictive models have been developed, few have been implemented prospectively. An important step towards implementation is building the informatics infrastructure for score calculation and assessing performance during silent implementation. During prospective validation, we found that eCART had similar performance to the retrospective evaluation. These results are encouraging, given that data quality and timing can differ between prospective and retrospective data. The head-to-head comparison of retrospectively calculated eCART scores in the prospective cohort to the scores filed back to the EHR in real time demonstrated a 12% increase in number of scores as well as a slightly higher average score, associated with a slightly lower AUROC. There are several potential causes for this discrepancy, including the correction of previously filed values and backdating of vital signs. Nevertheless, these analyses increase confidence that eCART will continue to perform well during implementation studies. Furthermore, we demonstrate the feasibility of testing these models in environments for clinical operations integrated with HL7.

Our study has several limitations. First, GBM models are complex and difficult to interpret. Therefore, explainable machine learning approaches are needed to provide insights to clinicians regarding the variables that are driving an individual patient’s risk of deterioration. In addition, there are myriad other machine learning approaches available to develop prediction models, and numerous possible comparator scores that have been published. We chose GBM due to its excellent discrimination and calibration from prior publications, and NEWS and MEWS due to their widespread use across the country and around the world. Finally, it is also important to note that high model discrimination may not translate to improved patient outcomes, so prospective implementation of eCART is required to study its impact on patient care.

## CONCLUSIONS

In conclusion, we developed and validated a new GBM model, eCARTv5, which accurately identifies early clinical deterioration. Our model was validated retrospectively in a geographically diverse set of health systems and performed better than the NEWS and MEWS across multiple subgroups. Prospective validation of eCART found similar performance, and these results served as the foundation for an FDA 510(k) clearance. Future implementation of our score could identify more high-risk patients at a lower false alarm rate than commonly used tools.

## Data Availability

The data utilized in this article cannot be shared publicly because of legal and regulatory restrictions. These data were obtained from four hospital systems after our research protocol was reviewed by IRBs from each hospital, and our data use agreements do not permit sharing due to the granular nature of the data.

## ACKNOWLEDGEMENTS

None

## Author contributions

MMC takes full responsibility for the content of the manuscript. MMC and DPE conceptualized the study. KAC conducted the statistical analysis of the data. MMC wrote the first draft of the manuscript and revised subsequent versions. All authors contributed to the interpretation of data, reviewed and edited the initial drafts, and approved the final manuscript.

## Financial support used for the study

This work was supported by funding from the National Institutes of Health (PI: MMC; R01HL157262) and Biomedical Advanced Research and Development Authority (BARDA) as part of its Division of Research Innovation and Ventures (DRIVe) under contract number 75A50121C00043 (PI: DPE).

## Disclosures and conflicts of interest

Drs. Churpek and Edelson are inventors on a patent for patient risk evaluation (US11410777) and receive royalties from this intellectual property from the University of Chicago. Dr. Edelson is employed by and has an equity stake in AgileMD, which markets and distributes eCART.

## Notes

### Funding Statement

This study was funded by the National Institutes of Health (PI: MMC; R01HL157262) and Biomedical Advanced Research and Development Authority (BARDA) as part of its Division of Research Innovation and Ventures (DRIVe) under contract number 75A50121C00043 (PI: DPE).

### Author Declarations

The following Institutional Review Boards (IRB) gave ethical approval for this work: University of Chicago Biological Sciences Division IRB (#18-0447), Loyola University Chicago Health Sciences Division IRB (#215437), NorthShore University HealthSystem Research Institute IRB (#EH16-210T), University of Wisconsin-Madison Minimal Risk Research IRB (#2019-1258), BayCare Health System IRB (#2022.014-B.MPH & #2022.015-B.MPH) and Yale Human Research Protection Program IRBs (#2000035317).

### Summary of Updates

Title change, only in PDF Manuscript and Online Supplement

