## Supplementary material for "Multicenter Development and Prospective Validation of eCARTv5: A Gradient Boosted Machine Learning Early Warning Score": ONLINE DATA SUPPLEMENT

**Corresponding author:**

Matthew M Churpek, MD, MPH, PhD

**ONLINE DATA SUPPLEMENT**

**Table of Contents:**

| **Supplement Section** | **Page** |
| --- | --- |
| **Supplemental Methods**  Additional Institutional Review Board Information  Model Development  Subgroup Definitions | **3**  3  4  4 |
| **Table E1.** Variables included in the eCART model. | **6** |
| **Table E2.** Non-physiologic filters for predictor variables. | **7** |
| **Table E3.** Heatmap distribution of variable missingness at the level of each eCART score for the derivation health system cohort. | **8** |
| **Table E4.** Heatmap distribution of variable missingness at the level of each eCART score for the retrospective and prospective validation cohorts by health system cohorts. | **12** |
| **Table E5.** Full retrospective cohort test characteristics for the primary outcome of deterioration [N=1,769,461 encounters]. | **16** |
| **Table E6.** Full retrospective cohort NEWS test characteristics for the primary outcome of deterioration [N=1,769,461 encounters]. | **17** |
| **Table E7.** Full retrospective cohort MEWS test characteristics for the primary outcome of deterioration [N=1,769,461 encounters]. | **18** |
| **Figure E1.** eCART partial plots. Partial dependence plots of the association between maximum respiratory rate in the prior 24 hours (A), delivered FiO2 (B), minimum systolic blood pressure in the prior 24 hours, and heart rate (D) and the risk of the outcome. | **19** |

**Supplemental Methods**

**Additional Institutional Review Board Information**

The study was approved by the following Institutional Review Boards (IRB) with a waiver of informed consent:

1. The University of Chicago Biological Sciences Division IRB (#18-0447; Title: Epidemiology and prediction of critical care interventions during rapid response team calls; Initial approval 6/20/2018; relevant amendment approved 10/8/2021.)
2. Loyola University Chicago Health Sciences Division IRB (#215437; Title: “Developing a clinical decision support tool for the identification, diagnosis, and treatment of critical illness in hospitalized patients”; Initial approval 12/17/2021; relevant amendments approved 03/07/2022 and 8/31/2022.)
3. NorthShore University HealthSystem Research Institute IRB (#EH16-210T; Title: “Sepsis Early Prediction and Subphenotype Illumination Study”; Initial approval 08/02/2016; relevant amendment approved 04/19/2021.)
4. The University of Wisconsin-Madison Minimal Risk Research IRB (#2019-1258; Title: “Predicting in-hospital clinical deterioration”; Initial approval 11/15/2019; relevant amendment approved 2/22/2021.)
5. BayCare Health System IRB
   a. (Retrospective Study: #2022.014-B.MPH; Title: “Validation of the eCART Score for Predicting Clinical Deterioration in Hospitalized Patients”; Initial Approval 2/23/2022)
   b. (Prospective Study: #2022.015-B.MPH; Title: ”A Rapid Diagnostic of Risk in Hospitalized Patients with COVID-19, Sepsis, and Other High-Risk Conditions to Improve Outcomes and Critical Resource Allocation Using Machine Learning”; Initial approval 2/23/2022)
6. Yale Human Research Protection Program IRBs (#2000035317; Title: “Validation of the electronic cardiac arrest risk triage (ECART) score for predicting clinical deterioration in hospitalized adults”; Initial approval 6/2/2023.)

**Model Development**

A gradient boosted machine (GBM) model was developed to predict clinical deterioration in all adult patients hospitalized on the wards in the training data. GBM models are ensembles of decision trees where each tree is iteratively built to improve upon the errors of the previous trees. In order to avoid overfitting and to optimize the final model, hyperparameters were tuned in the training cohort using five-fold cross-validation to maximize the area under the receiver operating characteristic curve (AUROC). Specifically, the number of trees (1000, 1500, 2000), interaction depth (20, 30, 40), shrinkage (0.001, 0.01, 0.1), and the minimum observations in a node (2, 4) were optimized, and the combination with the highest cross-validation AUROC was chosen as the final model.

**Subgroup Definitions**

Subgroup analyses were performed in the retrospective validation cohort across patient demographics (age, sex, race) and clinical conditions (surgical, obstetric, sepsis, COVID-19, congestive heart failure (CHF), and chronic obstructive pulmonary disease (COPD). Surgical patients were identified using hospital location data denoting an operating room location. Sepsis was defined using the suspicion of infection criteria published by Seymour et al. Specifically, a patient was identified as infected (“sepsis”) based on culture and antibiotic orders if either 1) a body fluid culture was ordered within 24 hours of an antibiotic if the antibiotic order was first or 2) the patient received an antibiotic in the next 72 hours if the body fluid culture order was first. COVID-19 was defined as either a positive COVID-19 test (PCR or antigen) or a COVID-19 diagnosis code during the admission. Obstetric patients were identified using diagnosis codes for birth or C-section in the current encounter, while COPD and CHF was defined using the Elixhauser criteria for these conditions based on diagnosis codes from the hospital encounter.

**Table E1.** Variables included in the eCART model.

| Demographics | Age | Continuous |
| --- | --- | --- |
| Vital signs | Temperature (C°), Heart Rate, Respiratory Rate, Systolic Blood Pressure (SBP), Diastolic Blood Pressure (DBP), O2 Saturation, Fraction of Inspired Oxygen (FiO2), AVPU, Disorientation (yes/no) | Continuous |
| Vital sign trends | Highest value in the last 24 hours of Heart Rate, Respiratory Rate, SBP, DBP, FiO2, AVPU (U=highest), and Disorientation (yes=highest) | Continuous |
|  | Lowest value in last 24 hours of Heart Rate, SBP, DBP, O2 Saturation, and FiO2 |  |
|  | Mean over the last 24 hours of Heart rate, Respiratory Rate, SBP, O2 Saturation, and FiO2 |  |
|  | Standard Deviation over the last 24 hours of Temperature, Heart Rate, Respiratory Rate, SBP, DBP, and FiO2 |  |
|  | Slope over the last 24 hours of Heart Rate, Respiratory Rate, Temperature, SBP, DBP, and FiO2 |  |
| Laboratory values | Basic Metabolic Panel [BMP]: Sodium, Chloride, Potassium, Bicarbonate (CO2), Anion Gap, Glucose, Calcium, Blood Urea Nitrogen (BUN), Serum Creatinine (SCr), Phosphate | Continuous |
|  | Liver Function Test [LFT]: Total Protein, Albumin, Total Bilirubin, Aspartate Aminotransferase (AST/SGOT), Alkaline Phosphatase |  |
|  | Complete Blood Count [CBC]: White Blood Cells (WBC), Hemoglobin, Platelet Count, Bands, Eosinophils, Lymphocytes, Monocytes, Neutrophils |  |
|  | Blood Gas test: Arterial pH, Venous pH, Arterial Partial Pressure of Oxygen, Arterial Partial Pressure of Carbon Dioxide, Venous Partial Pressure of Carbon Dioxide |  |
|  | Other labs: Lactate, Magnesium, Lipase, International Normalized Ratio (INR), Mean Corpuscular Volume (MCV), Partial Thromboplastin Time (PTT), and Red Cell Distribution Width (RDW) |  |
| Laboratory value trends | Change from last collected value of Potassium, SCr, BUN, CO2, Anion Gap, Glucose, Phosphate, WBC, Hemoglobin, and Platelet Count | Continuous |
| Nurse Documentation | Braden Scale (Activity, Friction and Shear, Mobility, Moisture, Nutrition, Sensory Perception, Total Score), Body Mass Index, Change from last recorded BMI | Continuous |
| Location | Prior Intensive Care Unit stay | Binary |
| Length of stay | Hours since admission until the current time point | Continuous |
| Time of Day | Hours since midnight of the current day | Continuous |
| Urinary Output | Sum of Urine Output over the last 24 hours | Continuous |

Abbreviations: eCART = electronic Cardiac Arrest Risk Triage score; AVPU = Alert, responds to Voice, responds to Pain, Unresponsive

**Table E2.** Non-physiologic filters for predictor variables.

| **Variable** | **Filter threshold to change to missing** |
| --- | --- |
| Temperature, °C | < 32 or > 44 |
| Respiratory rate, breaths per minute | < 1 or > 70 |
| Heart rate, beats per minute | < 1 or > 300 |
| Systolic blood pressure, mm Hg | < 30 or > 300 |
| Diastolic blood pressure, mm Hg | < 1 or > 250 |
| Peripheral oxygen saturation, % | < 11 or > 100 |
| FiO2 delivered, % | <21 or >100 |
| White blood cells, x10^9^/L | > 1000 |
| White blood cell components, % | <0 or >100 |
| Body mass Index, kg/m^2^ | < 10 or > 200 |

**Table E3.** Heatmap distribution of variable missingness at the level of each eCART score for the derivation health system cohort.

| **eCART variable** | **Derivation Cohort** | | |
| --- | --- | --- | --- |
|  | **D1** | **D2** | **D3** |
| Temperature | 0.4% | 0.5% | 0.9% |
| Temperature, std dev | 2.7% | 3.4% | 6.0% |
| Temperature, slope | 2.7% | 3.4% | 6.0% |
| Heart Rate | 0.5% | 0.4% | 0.4% |
| Heart Rate, Maximum | 3.0% | 3.2% | 4.3% |
| Heart Rate, Mean | 3.0% | 3.2% | 4.3% |
| Heart Rate, Minimum | 3.0% | 3.2% | 4.3% |
| Heart Rate, std dev | 3.0% | 3.2% | 4.3% |
| Heart Rate, slope | 3.0% | 3.2% | 4.3% |
| Systolic Blood Pressure | 0.3% | 0.4% | 1.0% |
| Systolic Blood Pressure, Max | 2.8% | 3.2% | 5.6% |
| Systolic Blood Pressure, Mean | 2.8% | 3.2% | 5.6% |
| Systolic Blood Pressure, Min | 2.8% | 3.2% | 5.6% |
| Systolic Blood Pressure, Std dev | 2.8% | 3.2% | 5.6% |
| Systolic Blood Pressure, Slope | 2.8% | 3.2% | 5.6% |
| Diastolic Blood Pressure | 0.3% | 0.4% | 1.0% |
| Diastolic Blood Pressure, Max | 2.8% | 3.2% | 5.6% |
| Diastolic Blood Pressure, Min | 2.8% | 3.2% | 5.6% |
| Diastolic Blood Pressure, Std dev | 2.8% | 3.2% | 5.6% |
| Diastolic Blood Pressure, Slope | 2.8% | 3.2% | 5.6% |
| Respirations | 0.4% | 0.4% | 0.8% |
| Respirations, Max | 2.5% | 3.2% | 5.3% |
| Respirations, Mean | 2.5% | 3.2% | 5.3% |
| Respirations, Std dev | 2.5% | 3.2% | 5.3% |
| Respirations, Slope | 2.5% | 3.2% | 5.3% |
| Oxygen Saturation | 0.5% | 0.8% | 2.1% |
| Oxygen Saturation, Min | 2.8% | 6.4% | 14.9% |
| Oxygen Saturation, Mean | 2.8% | 6.4% | 14.9% |
| FiO2 | 3.1% | 6.0% | 29.2% |
| FiO2, Max | 10.6% | 18.4% | 47.0% |
| FiO2, Mean | 10.6% | 18.4% | 47.0% |
| FiO2, Min | 10.6% | 18.4% | 47.0% |
| FiO2, Std div | 10.6% | 18.4% | 47.0% |
| FiO2, Slope | 10.6% | 18.4% | 47.0% |
| BMI | 10.1% | 61.3% | 45.1% |
| BMI, delta | 37.9% | 88.1% | 74.2% |
| Urine Output | 0.0% | 0.0% | 0.0% |
| AVPU | 6.1% | 13.4% | 1.9% |
| AVPU, Least responsiveness (24hrs) | 13.2% | 68.1% | 7.4% |
| AVPU, Disoriented | 7.0% | 11.3% | 2.7% |
| AVPU, Ever disoriented (24hrs) | 16.2% | 44.8% | 9.1% |
| Braden Scale | 3.5% | 12.9% | 6.0% |
| Braden Scale, Sensory | 3.4% | 5.0% | 5.9% |
| Braden Scale, Moisture | 3.5% | 5.0% | 5.9% |
| Braden Scale, Activity | 3.4% | 5.1% | 5.9% |
| Braden Scale, Mobility | 3.4% | 5.0% | 5.9% |
| Braden Scale, Nutrition | 3.4% | 5.0% | 5.9% |
| Braden Scale, Friction | 3.5% | 5.1% | 6.0% |
| White Blood Cells | 5.5% | 8.2% | 7.4% |
| White Blood Cells, Delta | 21.1% | 26.9% | 21.6% |
| White Blood Cells, Neutrophils | 39.6% | 28.1% | 74.7% |
| White Blood Cells, Bands | 65.9% | 26.3% | 26.7% |
| White Blood Cells, Lymphocytes | 30.2% | 32.4% | 26.9% |
| White Blood Cells, Monocytes | 30.3% | 25.2% | 27.1% |
| White Blood Cells, Eosinophils | 35.0% | 27.1% | 28.7% |
| Hemoglobin | 5.4% | 7.8% | 6.1% |
| Hemoglobin, Delta | 19.4% | 26.1% | 21.8% |
| Hemoglobin, MCV | 5.5% | 8.2% | 6.8% |
| Hemoglobin, RDW | 5.6% | 8.2% | 6.8% |
| Platelets | 5.5% | 8.2% | 7.8% |
| Platelets, Delta | 21.0% | 27.0% | 25.1% |
| aPTT | 51.8% | 69.6% | 51.7% |
| INR | 38.1% | 51.7% | 40.1% |
| Glucose | 8.3% | 17.4% | 9.7% |
| Glucose, Delta | 21.0% | 36.0% | 24.2% |
| Sodium | 8.3% | 17.5% | 9.7% |
| Potassium | 8.4% | 17.4% | 9.7% |
| Potassium, Delta | 21.2% | 35.9% | 24.2% |
| Chloride | 8.3% | 17.5% | 9.7% |
| Carbon Dioxide | 8.3% | 17.2% | 8.2% |
| Carbon Dioxide, Delta | 21.1% | 35.3% | 22.8% |
| Anion Gap | 8.3% | 17.5% | 9.7% |
| Anion Gap, Delta | 21.1% | 36.0% | 24.3% |
| BUN | 8.3% | 17.3% | 10.1% |
| BUN, Delta | 21.0% | 36.0% | 25.0% |
| Creatinine | 8.3% | 17.2% | 9.9% |
| Creatinine, Delta | 21.0% | 35.9% | 24.9% |
| Calcium | 8.2% | 17.4% | 10.3% |
| Magnesium | 18.1% | 63.9% | 32.1% |
| Phosphate | 19.6% | 74.9% | 37.9% |
| Phosphate, Delta | 34.3% | 84.2% | 50.6% |
| Protein | 34.8% | 54.8% | 41.9% |
| Albumin | 34.2% | 53.3% | 41.0% |
| Bilirubin, Total | 34.8% | 55.0% | 41.9% |
| Alk Phos | 34.9% | 55.0% | 42.0% |
| AST / SGOT | 35.0% | 54.8% | 41.9% |
| Lactate | 94.5% | 97.3% | 95.9% |
| Lipase | 81.9% | 83.9% | 85.7% |
| ABG pH | 98.8% | 98.7% | 87.3% |
| ABG pCO2 | 98.7% | 98.7% | 98.3% |
| ABG pO2 | 98.7% | 98.7% | 98.3% |
| VBG pH | 98.2% | 99.8% | 99.7% |
| VBG pCO2 | 98.2% | 99.8% | 99.7% |
| Age on admission | 0.0% | 0.0% | 0.0% |
| Length of stay | 0.0% | 0.0% | 0.0% |
| ICU this admission | 0.0% | 0.0% | 0.0% |
| Time of day | 0.0% | 0.0% | 0.0% |

Abbreviations: eCART = electronic Cardiac Arrest Risk Triage score

**Table E4.** Heatmap distribution of variable missingness at the level of each eCART score for the retrospective and prospective validation cohorts by health system cohorts.

| **eCART variable** | **Health System 1** | | **Health System 2** | | **Health System 3** | | |
| --- | --- | --- | --- | --- | --- | --- | --- |
|  | **Retrospective, R1** | **Prospective,**  **P1** | **Retrospective, R2** | **Prospective, P2** | **Retrospective,**  **R3** | **Prospective (Real Time Score), P3** | **Retrospectively Calculated Prospective, P3(r)** |
| Temperature | 0.2% | 0.4% | 0.1% | 0.1% | 0.1% | 0.2% | 0.1% |
| Temperature, std dev | 4.3% | 3.9% | 1.6% | 2.6% | 1.9% | 3.6% | 1.9% |
| Temperature, slope | 4.3% | 3.9% | 1.6% | 2.6% | 1.9% | 3.6% | 1.9% |
| Heart Rate | 0.1% | 0.0% | 0.1% | 0.0% | 0.1% | 0.0% | 0.1% |
| Heart Rate, Maximum | 2.6% | 1.4% | 1.0% | 1.4% | 1.0% | 0.7% | 0.7% |
| Heart Rate, Mean | 2.6% | 1.4% | 1.0% | 1.4% | 1.0% | 0.7% | 0.7% |
| Heart Rate, Minimum | 2.6% | 1.4% | 1.0% | 1.4% | 1.0% | 0.7% | 0.7% |
| Heart Rate, std dev | 2.6% | 1.4% | 1.0% | 1.4% | 1.0% | 0.7% | 0.7% |
| Heart Rate, slope | 2.6% | 1.4% | 1.0% | 1.4% | 1.0% | 0.7% | 0.7% |
| Systolic Blood Pressure | 0.2% | 0.0% | 0.2% | 0.1% | 0.0% | 0.0% | 0.0% |
| Systolic Blood Pressure, Max | 3.3% | 1.6% | 1.6% | 2.0% | 0.6% | 0.6% | 0.5% |
| Systolic Blood Pressure, Mean | 3.3% | 1.6% | 1.6% | 2.0% | 0.6% | 0.6% | 0.5% |
| Systolic Blood Pressure, Min | 3.3% | 1.6% | 1.6% | 2.0% | 0.6% | 0.6% | 0.5% |
| Systolic Blood Pressure, Std dev | 3.3% | 1.6% | 1.6% | 2.0% | 0.6% | 0.6% | 0.5% |
| Systolic Blood Pressure, Slope | 3.3% | 1.6% | 1.6% | 2.0% | 0.6% | 0.6% | 0.5% |
| Diastolic Blood Pressure | 0.2% | 0.0% | 0.2% | 0.1% | 0.0% | 0.0% | 0.0% |
| Diastolic Blood Pressure, Max | 3.3% | 1.6% | 1.6% | 2.0% | 0.6% | 0.6% | 0.5% |
| Diastolic Blood Pressure, Min | 3.3% | 1.6% | 1.6% | 2.0% | 0.6% | 0.6% | 0.5% |
| Diastolic Blood Pressure, Std dev | 3.3% | 1.6% | 1.6% | 2.0% | 0.6% | 0.6% | 0.5% |
| Diastolic Blood Pressure, Slope | 3.3% | 1.6% | 1.6% | 2.0% | 0.6% | 0.6% | 0.5% |
| Respirations | 0.3% | 0.4% | 0.1% | 0.1% | 0.2% | 0.3% | 0.3% |
| Respirations, Max | 2.8% | 2.5% | 1.4% | 2.2% | 7.4% | 11.3% | 11.0% |
| Respirations, Mean | 2.8% | 2.5% | 1.4% | 2.2% | 7.4% | 11.3% | 11.0% |
| Respirations, Std dev | 2.8% | 2.5% | 1.4% | 2.2% | 7.4% | 11.3% | 11.0% |
| Respirations, Slope | 2.8% | 2.5% | 1.4% | 2.2% | 7.4% | 11.3% | 11.0% |
| Oxygen Saturation | 0.4% | 0.1% | 0.2% | 0.1% | 0.2% | 0.1% | 0.1% |
| Oxygen Saturation, Min | 4.4% | 1.6% | 1.3% | 1.6% | 1.6% | 0.9% | 0.9% |
| Oxygen Saturation, Mean | 4.4% | 1.6% | 1.3% | 1.6% | 1.6% | 0.9% | 0.9% |
| FiO2 | 0.9% | 1.0% | 1.3% | 1.2% | 0.2% | 0.8% | 0.2% |
| FiO2, Max | 8.6% | 8.6% | 7.7% | 8.6% | 4.5% | 5.4% | 2.4% |
| FiO2, Mean | 8.6% | 8.6% | 7.7% | 8.6% | 4.5% | 5.4% | 2.4% |
| FiO2, Min | 8.6% | 8.6% | 7.7% | 8.6% | 4.5% | 5.4% | 2.4% |
| FiO2, Std div | 8.6% | 8.6% | 7.7% | 8.6% | 4.5% | 5.4% | 2.4% |
| FiO2, Slope | 8.6% | 8.6% | 7.7% | 8.7% | 4.5% | 5.4% | 2.4% |
| BMI | 4.3% | 4.4% | 10.8% | 15.8% | 11.6% | 8.5% | 8.3% |
| BMI, delta | 38.1% | 21.0% | 44.8% | 56.4% | 46.5% | 11.3% | 39.7% |
| Urine Output | 0.0% | 0.0% | 0.0% | 0.0% | 0.0% | 0.0% | 0.0% |
| AVPU | 1.4% | 12.7% | 2.7% | 6.2% | 0.3% | 0.3% | 0.1% |
| AVPU, Least responsiveness (24hrs) | 8.2% | 44.3% | 19.6% | 33.2% | 4.8% | 11.8% | 3.3% |
| AVPU, Disoriented | 8.9% | 3.2% | 1.4% | 40.1% | 0.7% | 9.7% | 0.4% |
| AVPU, Ever disoriented (24hrs) | 20.5% | 9.1% | 16.3% | 67.9% | 8.8% | 49.6% | 7.7% |
| Braden Scale | 2.1% | 5.2% | 1.6% | 6.3% | 6.2% | 6.4% | 3.5% |
| Braden Scale, Sensory | 2.1% | 5.2% | 1.5% | 6.2% | 5.9% | 6.4% | 3.5% |
| Braden Scale, Moisture | 1.9% | 5.2% | 1.5% | 6.3% | 5.9% | 6.4% | 3.5% |
| Braden Scale, Activity | 1.6% | 5.2% | 1.5% | 6.3% | 5.9% | 6.4% | 3.5% |
| Braden Scale, Mobility | 2.1% | 5.2% | 1.5% | 6.3% | 5.9% | 6.4% | 3.5% |
| Braden Scale, Nutrition | 2.1% | 5.2% | 1.5% | 6.3% | 5.9% | 6.4% | 3.5% |
| Braden Scale, Friction | 2.1% | 5.2% | 1.5% | 6.7% | 5.9% | 6.4% | 3.5% |
| White Blood Cells | 10.9% | 5.7% | 11.1% | 3.5% | 4.1% | 8.8% | 2.0% |
| White Blood Cells, Delta | 31.2% | 23.8% | 22.6% | 19.1% | 28.9% | 33.8% | 24.7% |
| White Blood Cells, Neutrophils | 25.3% | 25.2% | 2.9% | 6.5% | 6.1% | 13.1% | 7.0% |
| White Blood Cells, Bands | 92.6% | 28.3% | 81.8% | 7.4% | 86.7% | 93.5% | 93.1% |
| White Blood Cells, Lymphocytes | 25.3% | 25.2% | 4.2% | 6.5% | 6.1% | 13.2% | 7.1% |
| White Blood Cells, Monocytes | 25.3% | 25.2% | 4.2% | 6.5% | 6.2% | 13.3% | 7.2% |
| White Blood Cells, Eosinophils | 25.3% | 25.2% | 4.3% | 6.6% | 10.1% | 19.2% | 12.3% |
| Hemoglobin | 10.4% | 4.8% | 2.3% | 3.4% | 3.5% | 8.5% | 1.7% |
| Hemoglobin, Delta | 30.3% | 20.1% | 15.8% | 18.8% | 27.3% | 32.5% | 22.5% |
| Hemoglobin, MCV | 12.0% | 5.7% | 2.5% | 3.5% | 4.1% | 8.9% | 2.0% |
| Hemoglobin, RDW | 12.0% | 5.8% | 2.9% | 3.6% | 4.1% | 8.9% | 2.0% |
| Platelets | 10.7% | 5.6% | 2.5% | 3.5% | 4.1% | 8.6% | 2.0% |
| Platelets, Delta | 31.3% | 23.0% | 16.1% | 19.1% | 28.9% | 33.7% | 24.7% |
| aPTT | 71.3% | 70.2% | 59.0% | 63.1% | 68.8% | 76.1% | 75.2% |
| INR | 39.3% | 40.2% | 45.4% | 49.3% | 48.5% | 54.8% | 52.7% |
| Glucose | 10.5% | 5.9% | 6.5% | 6.2% | 11.1% | 7.0% | 6.0% |
| Glucose, Delta | 28.7% | 20.5% | 16.6% | 20.3% | 33.9% | 31.7% | 27.0% |
| Sodium | 9.3% | 5.4% | 6.5% | 6.1% | 5.2% | 2.9% | 2.7% |
| Potassium | 8.6% | 5.1% | 6.6% | 6.3% | 5.2% | 3.0% | 2.7% |
| Potassium, Delta | 25.6% | 18.9% | 16.8% | 19.7% | 26.9% | 25.9% | 22.3% |
| Chloride | 9.5% | 5.5% | 6.7% | 6.1% | 5.2% | 4.4% | 2.7% |
| Carbon Dioxide | 9.4% | 5.4% | 6.6% | 6.0% | 11.0% | 6.8% | 5.7% |
| Carbon Dioxide, Delta | 26.8% | 18.5% | 15.7% | 17.8% | 33.5% | 30.5% | 25.7% |
| Anion Gap | 9.6% | 5.5% | 6.7% | 6.2% | 11.1% | 6.9% | 5.9% |
| Anion Gap, Delta | 28.0% | 21.6% | 17.5% | 19.7% | 33.8% | 32.0% | 26.5% |
| BUN | 10.2% | 6.4% | 6.4% | 6.0% | 5.2% | 4.4% | 2.7% |
| BUN, Delta | 28.8% | 22.1% | 16.7% | 19.5% | 27.0% | 28.2% | 22.5% |
| Creatinine | 9.1% | 4.9% | 6.5% | 6.1% | 11.1% | 7.0% | 6.0% |
| Creatinine, Delta | 27.2% | 19.6% | 17.7% | 20.2% | 33.7% | 31.5% | 26.9% |
| Calcium | 13.3% | 7.9% | 6.8% | 6.2% | 8.1% | 4.5% | 3.9% |
| Magnesium | 35.8% | 22.0% | 31.5% | 30.3% | 47.8% | 41.1% | 38.9% |
| Phosphate | 51.8% | 42.0% | 51.6% | 49.3% | 74.2% | 79.9% | 78.4% |
| Phosphate, Delta | 66.8% | 58.0% | 66.1% | 64.5% | 86.9% | 89.9% | 89.0% |
| Protein | 55.1% | 38.4% | 29.2% | 26.4% | 22.8% | 20.0% | 19.6% |
| Albumin | 48.7% | 35.0% | 27.3% | 24.6% | 22.6% | 19.7% | 19.2% |
| Bilirubin, Total | 45.7% | 34.3% | 25.8% | 24.5% | 22.8% | 20.0% | 19.5% |
| Alk Phos | 47.8% | 34.7% | 25.8% | 23.4% | 22.8% | 20.0% | 19.6% |
| AST / SGOT | 44.4% | 33.1% | 25.8% | 24.2% | 23.0% | 20.1% | 19.7% |
| Lactate | 95.0% | 90.9% | 92.6% | 90.9% | 94.0% | 89.7% | 92.1% |
| Lipase | 81.5% | 79.6% | 72.2% | 75.1% | 79.8% | 81.1% | 81.1% |
| ABG pH | 97.6% | 97.9% | 97.9% | 98.2% | 98.0% | 97.1% | 97.6% |
| ABG pCO2 | 97.6% | 97.9% | 97.6% | 97.3% | 98.0% | 97.2% | 97.5% |
| ABG pO2 | 97.6% | 97.9% | 97.9% | 98.2% | 98.0% | 97.2% | 97.6% |
| VBG pH | 98.3% | 94.8% | 97.0% | 98.9% | 99.8% | 99.2% | 99.4% |
| VBG pCO2 | 98.3% | 94.8% | 97.0% | 98.9% | 99.8% | 99.7% | 99.4% |
| Age on admission | 0.0% | 0.0% | 0.0% | 0.0% | 0.0% | 0.0% | 0.0% |
| Length of stay | 0.0% | 0.0% | 0.0% | 0.0% | 0.0% | 0.0% | 0.0% |
| ICU this admission | 0.0% | 0.0% | 0.0% | 0.0% | 0.0% | 0.0% | 0.0% |
| Time of day | 0.0% | 0.0% | 0.0% | 0.0% | 0.0% | 0.0% | 0.0% |

**Table E5**. Full retrospective cohort test characteristics for the primary outcome of deterioration [N=1,769,461 encounters].

| **eCART** | **Sensitivity** | **Specificity** | **Positive Predictive Value** | **Negative Predictive Value** |
| --- | --- | --- | --- | --- |
| 88 | 61.2% (61.1%, 61.2%) | 88.4% (88.4%, 88.4%) | 6.5% (6.5%, 6.5%) | 99.4% (99.4%, 99.4%) |
| 89 | 59.6% (59.5%, 59.7%) | 89.3% (89.3%, 89.3%) | 6.9% (6.9%, 6.9%) | 99.4% (99.4%, 99.4%) |
| 90 | 57.9% (57.9%, 58.0%) | 90.2% (90.2%, 90.2%) | 7.3% (7.3%, 7.3%) | 99.4% (99.4%, 99.4%) |
| 91 | 56.1% (56.0%, 56.2%) | 91.2% (91.2%, 91.2%) | 7.8% (7.8%, 7.8%) | 99.4% (99.4%, 99.4%) |
| 92 | 54.0% (54.0%, 54.1%) | 92.1% (92.1%, 92.1%) | 8.4% (8.3%, 8.4%) | 99.3% (99.3%, 99.3%) |
| 93 | 51.8% (51.7%, 51.8%) | 93.1% (93.1%, 93.1%) | 9.0% (9.0%, 9.1%) | 99.3% (99.3%, 99.3%) |
| 94 | 49.2% (49.1%, 49.3%) | 94.0% (94.0%, 94.0%) | 9.9% (9.8%, 9.9%) | 99.3% (99.3%, 99.3%) |
| 95 | 46.3% (46.2%, 46.4%) | 95.0% (95.0%, 95.0%) | 10.9% (10.9%, 10.9%) | 99.3% (99.3%, 99.3%) |
| 96 | 42.9% (42.8%, 42.9%) | 95.9% (95.9%, 95.9%) | 12.3% (12.2%, 12.3%) | 99.2% (99.2%, 99.2%) |
| 97 | 38.6% (38.5%, 38.7%) | 96.9% (96.9%, 96.9%) | 14.2% (14.1%, 14.2%) | 99.2% (99.2%, 99.2%) |
| 98 | 33.1% (33.0%, 33.2%) | 97.9% (97.9%, 97.9%) | 17.1% (17.0%, 17.1%) | 99.1% (99.1%, 99.1%) |
| 99 | 24.6% (24.5%, 24.7%) | 98.9% (98.9%, 98.9%) | 22.4% (22.3%, 22.4%) | 99.0% (99.0%, 99.0%) |
| 99.1 | 23.4% (23.4%, 23.5%) | 99.0% (99.0%, 99.0%) | 23.2% (23.1%, 23.3%) | 99.0% (99.0%, 99.0%) |
| 99.2 | 22.2% (22.1%, 22.3%) | 99.1% (99.1%, 99.1%) | 24.1% (24.1%, 24.2%) | 99.0% (99.0%, 99.0%) |
| 99.3 | 20.8% (20.8%, 20.9%) | 99.2% (99.2%, 99.2%) | 25.2% (25.1%, 25.3%) | 98.9% (98.9%, 99.0%) |

Abbreviations: UW = UW Health University Hospital; YNHHS = Yale New Haven Health System; eCART = electronic Cardiac Arrest Risk Triage score

**Table E6.** Full retrospective cohort NEWS test characteristics for the primary outcome of deterioration [N=1,769,461 encounters].

| **NEWS** | **Sensitivity** | **Specificity** | **Positive Predictive Value** | **Negative Predictive Value** |
| --- | --- | --- | --- | --- |
| 4 | 60.2% (60.1%, 60.3%) | 80.8% (80.8%, 80.8%) | 4.0% (4.0%, 4.0%) | 99.3% (99.3%, 99.4%) |
| 5 | 49.7% (49.6%, 49.8%) | 88.3% (88.3%, 88.3%) | 5.4% (5.3%, 5.4%) | 99.2% (99.2%, 99.3%) |
| 6 | 37.9% (37.8%, 37.9%) | 93.6% (93.6%, 93.6%) | 7.3% (7.3%, 7.3%) | 99.1% (99.1%, 99.1%) |
| 7 | 28.0% (27.9%, 28.1%) | 96.6% (96.6%, 96.6%) | 9.9% (9.8%, 9.9%) | 99.0% (99.0%, 99.0%) |
| 8 | 20.1% (20.1%, 20.2%) | 98.2% (98.2%, 98.2%) | 13.0% (12.9%, 13.0%) | 98.9% (98.9%, 98.9%) |
| 9 | 13.3% (13.3%, 13.4%) | 99.1% (99.1%, 99.1%) | 16.8% (16.7%, 16.8%) | 98.9% (98.8%, 98.9%) |

Abbreviations: UW = UW Health University Hospital; YNHHS = Yale New Haven Health System; NEWS = National Early Warning Score

**Table E7.** Full retrospective cohort MEWS test characteristics for the primary outcome of deterioration [N=1,769,461 encounters].

| **MEWS** | **Sensitivity** | **Specificity** | **Positive Predictive Value** | **Negative Predictive Value** |
| --- | --- | --- | --- | --- |
| 2 | 62.2% (62.1%, 62.2%) | 72.6% (72.6%, 72.6%) | 2.9% (2.9%, 2.9%) | 99.3% (99.3%, 99.3%) |
| 3 | 38.9% (38.8%, 39.0%) | 91.5% (91.5%, 91.5%) | 5.7% (5.7%, 5.7%) | 99.1% (99.1%, 99.1%) |
| 4 | 22.6% (22.5%, 22.7%) | 97.4% (97.4%, 97.4%) | 10.4% (10.4%, 10.5%) | 99.0% (99.0%, 99.0%) |
| 5 | 12.1% (12.0%, 12.1%) | 99.2% (99.2%, 99.2%) | 16.9% (16.9%, 17.0%) | 98.8% (98.8%, 98.8%) |

Abbreviations: UW = UW Health University Hospital; YNHHS = Yale New Haven Health System; MEWS = Modified Early Warning Score

**Figure E1. eCART partial plots.** Partial dependence plots of the association between maximum respiratory rate in the prior 24 hours (A), delivered FiO2 (B), minimum systolic blood pressure in the prior 24 hours, and heart rate (D) and the risk of the outcome.

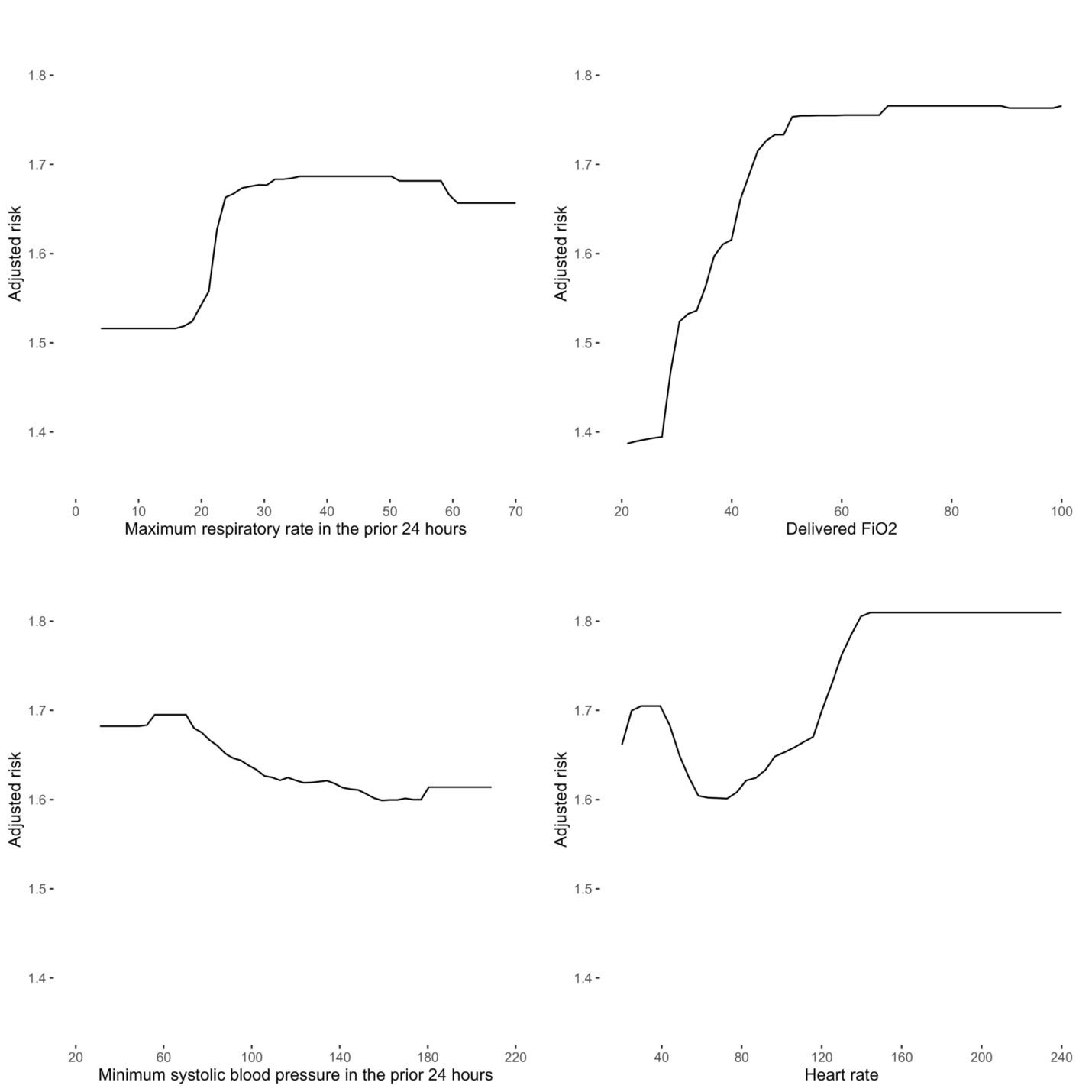

B.

A.

C.

D.

Abbreviations: eCART = electronic Cardiac Arrest Risk Triage score; FiO2 = Fraction of Inspired Oxygen
